# Precision Insulin-Pump Calibration and Optimal Insulin Delivery to Improve Blood Glucose Levels

**DOI:** 10.64898/2026.07.27.26359073

**Authors:** Iman Aganj, Nora Bryant, Leah (Morgan) Panaro, Peter Caravan, Jason L. Gaglia, Bruce Fischl

## Abstract

The successful management of type-1 diabetes (T1D) and insulin-dependent type-2 diabetes depends on the ability to accurately calibrate bolus and basal insulin doses, and minimize the time spent with high postprandial blood glucose (BG) levels while avoiding dangerously low hypoglycemic excursions. Precise calibration of the insulin pump helps with successful disease management; however, people with T1D may still experience prolonged, potentially damaging BG levels due to postprandial highs. Insulin pumps offer large, currently underexploited degrees of freedom in insulin delivery, which can have a dramatic impact on average postprandial BG levels. Building on existing models of the glucose-insulin system, we first propose a simple, automated, and individualized insulin-pump calibration system based on a series of measures taken before and a few hours after carbohydrate ingestion. We then modulate the shape of pre-meal insulin dosing to explicitly reduce postprandial BG levels while minimizing the likelihood of dangerously low BG. Such an optimal insulin delivery time course can potentially improve postprandial BG levels and rapidly bring BG to the target level. We evaluated our methods in 20 patients over 4 days, with BG levels sampled frequently.

## Introduction

A large portion of people with type-1 diabetes (T1D),^1, 2^ as well as some with insulin-dependent type-2 diabetes (ID-T2D), use insulin pumps for subcutaneous insulin delivery. Such pumps typically mimic the function of the healthy pancreas by administering doses in two forms: a relatively large *bolus* that is taken prior to mealtimes or to bring high blood glucose (BG) levels down to target, and a smaller and more constant *basal* that is given as a square wave of varying level throughout the day. Pumps can also use a combination of these two: *dual-wave* infusions that give an upfront bolus, followed by the remainder of the dose as an elevated basal rate to account for slowly metabolized carbs in foods high in fat and/or protein. Insulin pumps can be difficult to calibrate, though, particularly in young children, in whom frequent and extended fasting, which is by far the easiest way to accurately estimate basal insulin needs, is problematic, and growth spurts and other developmental changes cause the various parameters to change frequently. Even with fasting, setting time-varying basal insulin levels is complicated by the long time delays between insulin administration and its effects on BG levels.

The current standard of care in pump calibration uses summary statistics compiled across populations to set pump constants, largely based on body weight.^3-5^ However, not accounting for intrinsic variability across individuals can result in errors in the settings,^5^ potentially leading to a large negative impact on average BG levels. Thus, it is critical for quality of life and long-term health that pump parameters be set more accurately than is possible based on population models.^6-9^ Unfortunately, accurate calibration of insulin pumps is difficult because BG levels depend on multiple parameters, including long time constants. This motivates our first and main contribution: an optimal estimation procedure for accurately calibrating insulin pumps based on individual patient information. Accurate calibration would reduce the risk of developing damaging long-term sequelae such as neuropathy and would improve the quality of life of persons living with T1D or ID-T2D who could have greater confidence that their BG levels will remain within the desired range.

With the advent of continuous glucose monitoring (CGM)^10^ in interstitial fluid, there has been a research effort focused on developing closed-loop systems for delivering appropriate insulin dosages based on real-time BG readings.^11-24^ These systems have been shown to be effective in both inpatient^11-14^ and outpatient^15^ studies. Further increases in control can be obtained if a soluble stable form of glucagon^25^ can be synthesized, allowing the control systems to both “push” and “pull” on BG levels, potentially reducing average BG levels by 30 mg/dL.^19, 20^

Closed-loop control holds great long-term potential for managing T1D, freeing patients with T1D from the burden of constant monitoring and frequent insulin dosing. Given the great appeal of closed-loop systems, open-loop control (requiring manual management of insulin delivery) has received less attention, despite its potential to substantially decrease patient A1C levels even with simple procedures.^26, 27^ Open-loop systems can be implemented much more rapidly, without requiring advances in sensor technology or stable soluble glucagon. In fact, open-loop systems are the current standard of care in T1D, but there has been little effort devoted to using optimal control theory or modeling of insulin delivery to the bloodstream to shape the BG response through insulin dosing. In our second contribution, we seek to improve postprandial BG levels in subjects with T1D. Insulin doses are commonly given in two-component waveforms – a delta-function bolus and a square-wave over time. This ignores an important advantage that insulin pumps have over injections: the freedom to modulate the amplitude and scheduling of insulin delivery. More specifically, with the advent of sophisticated and accurate mathematical models of the insulin-glucose system in T1D,^28-30^ we derive insulin amplitudes and time courses that are explicitly designed to minimize hypo- and hyperglycemic excursions from the target BG. It is important to note that this is, in spirit, no different than standard current clinical advice, such as pre-bolusing before meals, which has been shown to significantly reduce postprandial BG levels,^31^ the use of dual-wave boluses to account for long-acting carbs, which has been shown to reduce A1C levels,^32^ as well as super-bolusing, which is effective in avoiding postprandial hyperglycemia.^33-35^

Here we propose to build a computational framework around the insight that pre-bolusing can reduce postprandial BG levels, with potential gains.

In the following, we describe our methodology in the “Materials and Methods” section, present experimental results and discuss them in the “Results” and “Discussion” sections, and conclude the paper with some remarks in the “Conclusion” section.

## Materials and Methods

We use a set of linked ordinary differential equations to capture the time-varying response of BG levels to carbohydrate intake, yielding a closed-form model of insulin delivery and response. We apply this model to automatically compute the insulin pump parameters that are currently set by trial and error in current standards of care, and also to leverage a currently unused degree of freedom in dosing with insulin pumps – the ability to take future basal insulin and deliver it immediately – to reduce postprandial high BG levels. We have acquired data to calibrate and validate our proposed algorithms.

### Mathematical model

We begin by mathematically modeling the temporal evolution of the plasma BG, *G*(*t*), and the insulin level in the interstitial fluid, *I*_*f*_(*t*), and in plasma, *I*_*p*_(*t*), given the time-varying carb intake, *c*(*t*), and the amount of insulin that is administered by the pump per unit of time, *I*(*t*). We propose to use the following variation on the two-compartment model^30, 36^ for *G*(*t*), which has been shown^26^ to accurately fit individual subject data:

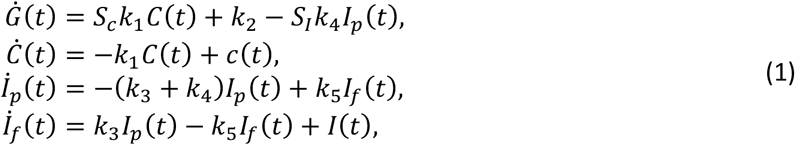

where *C*(*t*) is the amount of carbs not yet metabolized, *S*_*c*_ and *S*_*I*_ are positive constants representing the sensitivity of the BG to carbohydrates and insulin, respectively (with the insulin-to-carb ratio being 1:(*S*_*I*_⁄*S*_*C*_)), and *k*_1_, …, *k*_5_ are positive (inverse) time constants. The *dot* (·) above a variable denotes the temporal derivative. Assuming *C*_0_ carbs are ingested at time *t*_*c*_ ≥ 0, i.e. *c*(*t*) = *C*_0_*δ*(*t* − *t*_*c*_) with *δ*(·) the Dirac delta function, Eq. (1) leads to an exponential behavior for the remaining carbs, as:

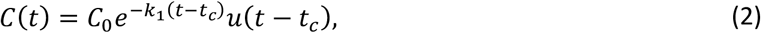

where *u*(·) is the Heaviside step function. The rate at which carbohydrates are metabolized, *k*_1_, is a food-specific parameter which is higher for rapid-acting carbs and lower for food with high fat and/or protein content. By parameterizing the problem with respect to *I*_*p*_, integrating *Ġ* leads to:

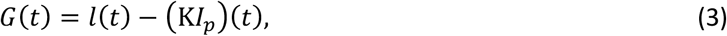

where K is an integral operator, and:

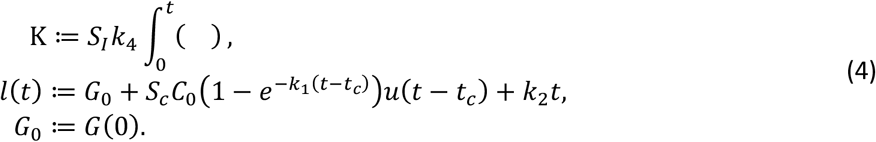

The plasma insulin can be solved from the administered insulin rate using the following second-order ordinary differential equation derived from Eq. (1):

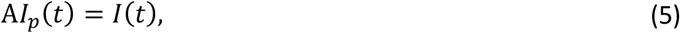

where the operator A is defined as:

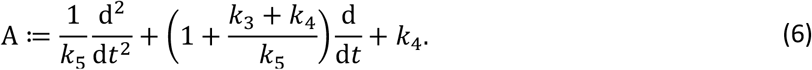

We conclude this subsection by focusing on some common forms of *I*, and providing the *particular* solution for *I*_*p*_ as well as for the term 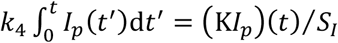 that would be substituted in Eq. (3) to calculate the BG over time.

- A constant unit basal, i.e. *I*(*t*) = 1, results in:

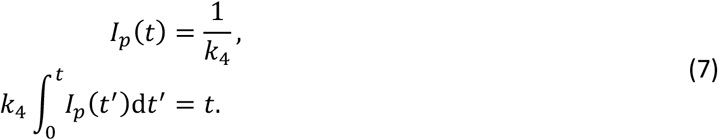
- A unit bolus at time *t*_*B*_ ≥ 0, i.e. *I*(*t*) = *δ*(*t* − *t*_*B*_), leads to:

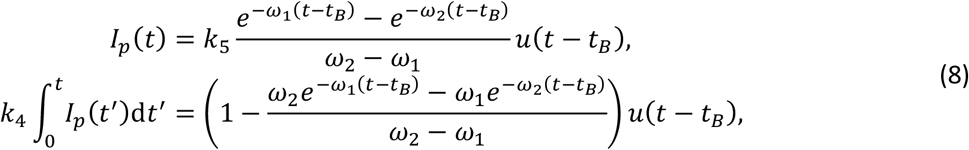

where the real-valued and positive *ω*_1_ < *ω*_2_ are:

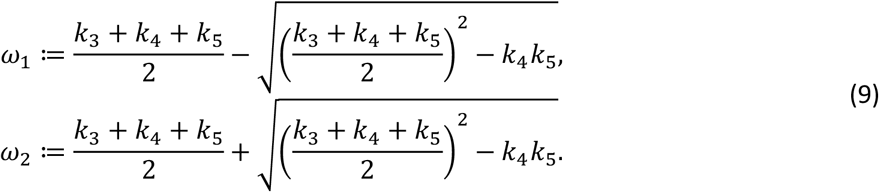
- A unit basal rate that starts at time *t*_*b*_ ≥ 0, i.e. *I*(*t*) = *u*(*t* − *t*_*b*_), results in:

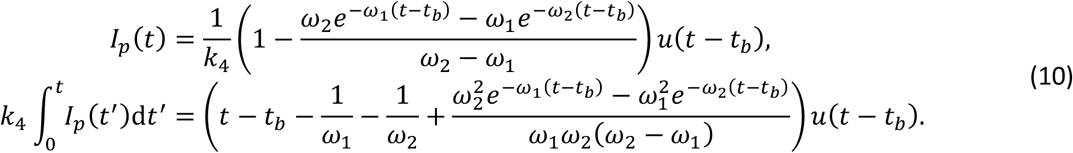

### Automated pump calibration

Using the aforementioned model, we propose automatically calibrating insulin pumps from individual BG measurements. Given a recent history of delivered insulin (boluses and basal rates) and ingested carbs by a patient, one can apply the above equations to solve the forward problem of predicting the temporal evolution of BG for any set of values for the individual’s physiological parameters, i.e., Θ ≔ {*S*_*I*_, *S*_*c*_, *k*_1_, …, *k*_5_}. However, we are rather interested in the *inverse* problem of estimating the patient-specific physiological parameters given a set of *N* BG measurements of an individual, *G*_0_, …, *G*_*N*−1_, at times *t*_0_, …, *t*_*N*−1_, which can help us to precisely calibrate the insulin pump for the patient. We do this by discretizing the space of the patient’s physiological parameters and exhaustively searching it for the values minimizing the following (robust L_*1*_) distance error between the predicted BG, *G*^Θ^(*t*_*n*_), and the measured BG, *G*_*n*_:

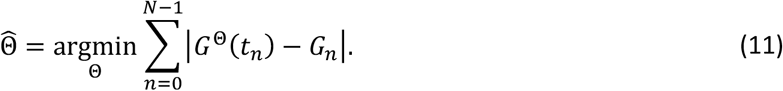

Given that an individual’s hepatic glucose rate, *k*_2_, is not necessarily constant throughout the day, we estimate two different values for it, along with the time of the day where *k*_2_ switches from its first value to the second. We also include the additional unknown parameter of carb ingestion delay, i.e., the difference between *t*_*c*_ and the mealtime, given its variability among subjects. Furthermore, since the values of *k*_3_, *k*_4_, and *k*_5_ cannot be uniquely determined from the estimated *ω*_1_ and *ω*_2_, we constrain *k*_4_ and *k*_5_ to be equal. In practice, such a constraint does not affect the subsequent optimal insulin scheduling (next subsection), given that the temporal dynamics of the glucose-insulin system will still be governed by *ω*_1_ and *ω*_2_ (that are unaffected by the constraint). In our experiments, therefore, Θ consisted of nine parameters, requiring us to search a nine-dimensional space. The optimization is highly parallelizable and can be considerably sped up using a graphics processing unit (GPU) and/or through a distributed search of the parameter space using a high-performance compute cluster. Assuming the consistency of the patient’s physiological parameters from day to day, Eq. (11) can be straightforwardly extended to the simultaneous optimization of parameters from measurements across multiple days by adding all the sums corresponding to different days into a single minimization.

### Insulin delivery optimization

The central insight in this step is that, in contrast to a healthy pancreas that delivers insulin directly into the bloodstream, insulin pumps deliver into subcutaneous tissue and hence can only gradually modify plasma insulin levels. While current pumps typically deliver insulin as a bolus and a square-wave basal, many unused degrees of freedom can be leveraged to reduce postprandial BG levels.

After estimating the patient’s physiological parameters 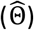 as in the previous subsection, we attempt here to optimize complex insulin waveforms to keep postprandial BG levels within the normal range as much as possible. We compute the optimal waveform of the delivered insulin for: a given amount of ingested carbs at a given time, the given initial BG level (*G*_0_), and a chosen target BG level (*G*_∞_). Our goal is to explicitly solve for a time-varying set of insulin doses, *Î*(*t*), during an interval 0 ≤ *t* ≤ *T* (before bringing it back to the estimated basal level of *k*_2_⁄*S*_*I*_), which minimizes the temporal average of the BG during the much longer interval 0 ≤ *t* ≤ *T*_∞_ while always keeping the BG above a safe level, *G*_*min*_. Mathematically:

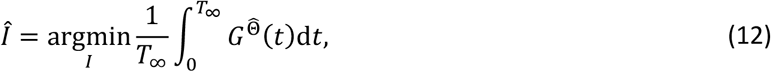

subject to:

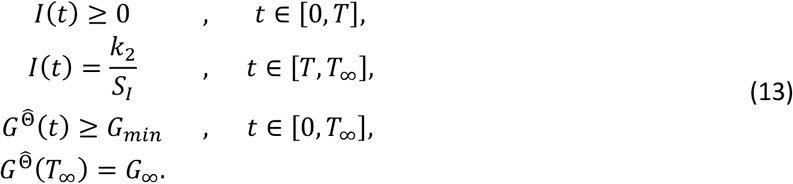

The first constraint, which keeps the insulin dose non-negative, could be removed to simulate the use of a bi-hormonal pump.^19, 20^ As in the previous subsections, we will parameterize the problem with respect to the plasma insulin, *I*_*p*_ ≥ 0. Equations (3), (5), and (12) result in the following maximization:

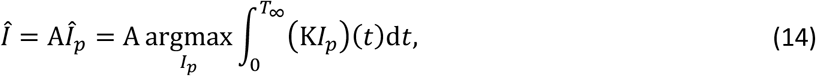

where the constraints are rewritten with respect to *I*_*p*_:

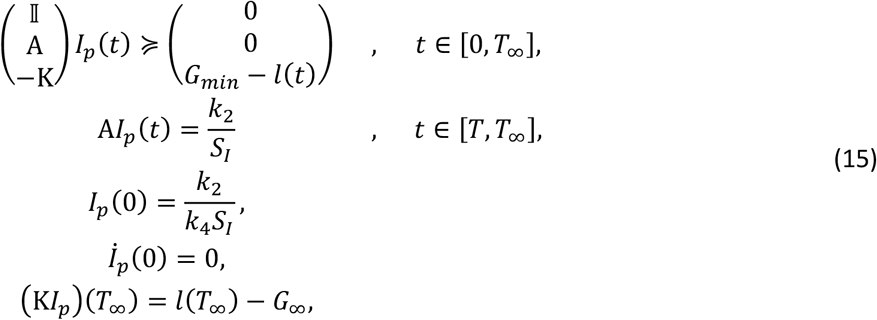

where I is the identity operator. The constraints on the initial values of *I*_*p*_ and its derivative ensure that the plasma insulin is initially stable and equal to the basal rate. By discretizing the values of *I*_*p*_ for a finite set of timepoints, Eq. (14) and the constraints (15) can all be seen to be linear with respect to the vectorized *I*_*p*_, meaning that the optimization problem is convex and can be efficiently solved via linear programming^37^.

### Data collection and processing

This study was approved by the Joslin Diabetes Center’s Committee on Human Studies. We acquired the data necessary to validate our models at the Joslin Diabetes Center. We enrolled 21 T1D patients and collected data from them during 4 visits (except for 2 subjects who dropped out after 2 visits and 1 visit, respectively), totaling 79 experiments. We excluded the patient who dropped out after 1 visit, keeping 20 subjects (10 females and 10 males) for our analysis.

At each visit, we measured intravenous BG levels every 10 minutes immediately before and for 4 hours after the administration of 33-66g of carbohydrates as part of a controlled mixed meal (Boost High Protein Drink). For the first half of our enrolled population, we prioritized using the target BG level recommended by the patient’s physician. For the patients in the second half of the population, however, we decided to keep the target BG to *G*_∞_ = 100 mg⁄dL for consistency. Since the target BG directly affects insulin scheduling, for each patient, we compare only visits with equal target BG values. To maximize the number of subjects included in our group analysis, we compare visits only in pairs (otherwise, if all visits were compared simultaneously, the constraint that visits have equal target BG would exclude more subjects).

In visit 1, we used a standard insulin schedule based on the patient’s insulin pump’s initial calibration. The standard schedule consisted of a bolus and a constant basal rate to counteract the carbs and the hepatic glucose, respectively, with the bolus being administered at the same time as the carbs. We then estimated model parameters from the data of visit 1 to be used for a standard schedule in visit 2. As previously described, our parameter-estimation algorithm used the BG values recorded during the experiment and the boluses administered in the past 12 hours, and exhaustively searched the space of the patient’s physiological parameters to find the values that best fit the data. Table 1 shows the range of values searched for each parameter. The first hypothesis we tested was that, for each subject, the mean BG was significantly lower and the target BG level was more precisely achieved in visit 2 than in visit 1. We then updated the model parameters using data from the first two visits via a more accurate joint estimation.

**Table 1.**
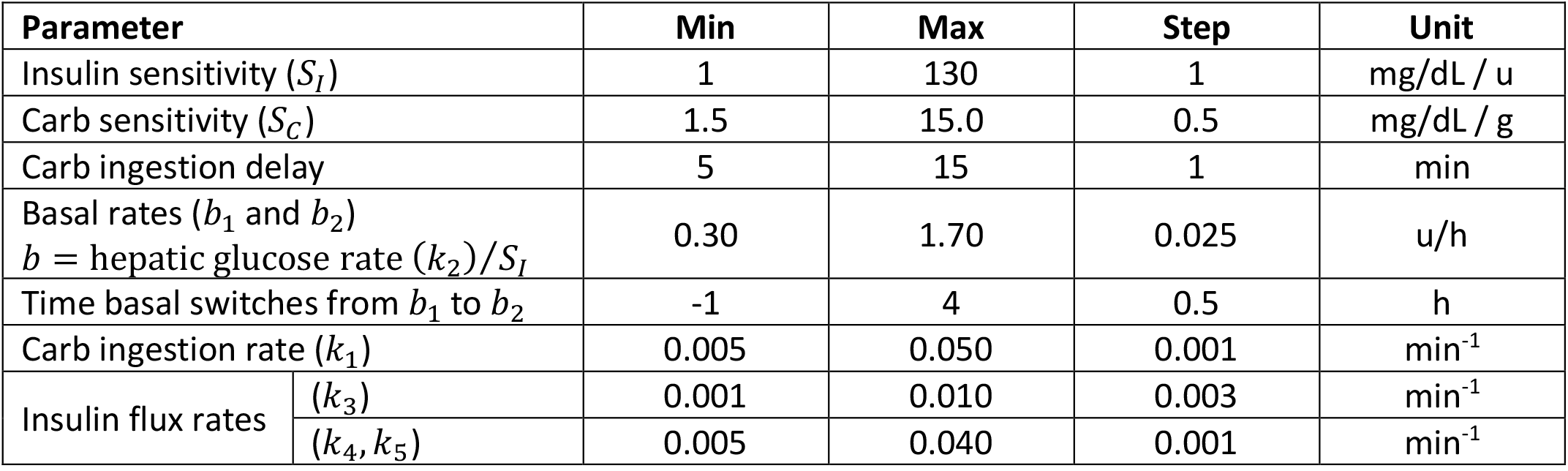
Range of the searched values for the patient’s physiological parameters.

Next, using the updated model parameters, we computed the optimal insulin delivery schedule as previously described for a series of initial BG levels. We employed linear programming (Matlab’s `linprog` function) to find an optimized *Î*_*p*_(*t*) and subsequently *Î*(*t*) (as discrete vectors with a time step size of 1 *min*) to suggest an optimal insulin schedule for a period of *T* = 2*h* given any initial BG value. We aimed to reduce the average BG over a period of *T*_∞_ = 5*T* = 10*h* while ensuring that the BG always stayed above *G*_*min*_ = 80 mg⁄dL, i.e. optimizing Eq. (14) subject to the constraints (15). We evaluated the schedules suggested by this algorithm by applying the schedule corresponding to the patient’s initial BG level in visit 3. We repeated this in visit 4 after making experimental refinements in the optimal schedule. For our last six patients, the consistent difference was that we allowed for pre-boluses as early as 35 min and 1h before the carb consumption in visits 3 and 4, respectively. The second hypothesis we tested was that for each subject, the mean BG was significantly lower (with BG safely above *G*_*min*_) and the target BG level was more precisely achieved using our optimal insulin schedule than the standard schedule.

## Results

### Pump calibration

For each of the visits 1 and 2 and each of the included 17 subjects (with equal target BG in the two visits), we computed average, minimum, and maximum BG during the ∼4h experiment, as well as the absolute distance of the final BG from the target BG and the overall reduction in BG. Table 2 shows the mean ± standard error of the mean (SEM) of these quantities, as well as two-tailed paired *t*-test *p*-values. As can be seen, all these quantities decreased from visit 1 (patient’s initial pump calibration) to visit 2 (calibration with our algorithm), and the reductions were almost always statistically significant. The evolution of the BG with time, averaged across subjects, is plotted for each visit in Figure 1. We calibrated the patient’s pump in visit 2 (by setting the basal values equal to the hepatic glucose rates estimated from visit 1) as soon as they arrived, i.e., up to half an hour to an hour before we started measuring the BG in visit 2. Consequently, the first recorded BG value was significantly lower in visit 2 than in visit 1. Nonetheless, the reduction in overall BG in visit 2 was larger than the difference in the initial BG between the two visits.

**Table 2.**
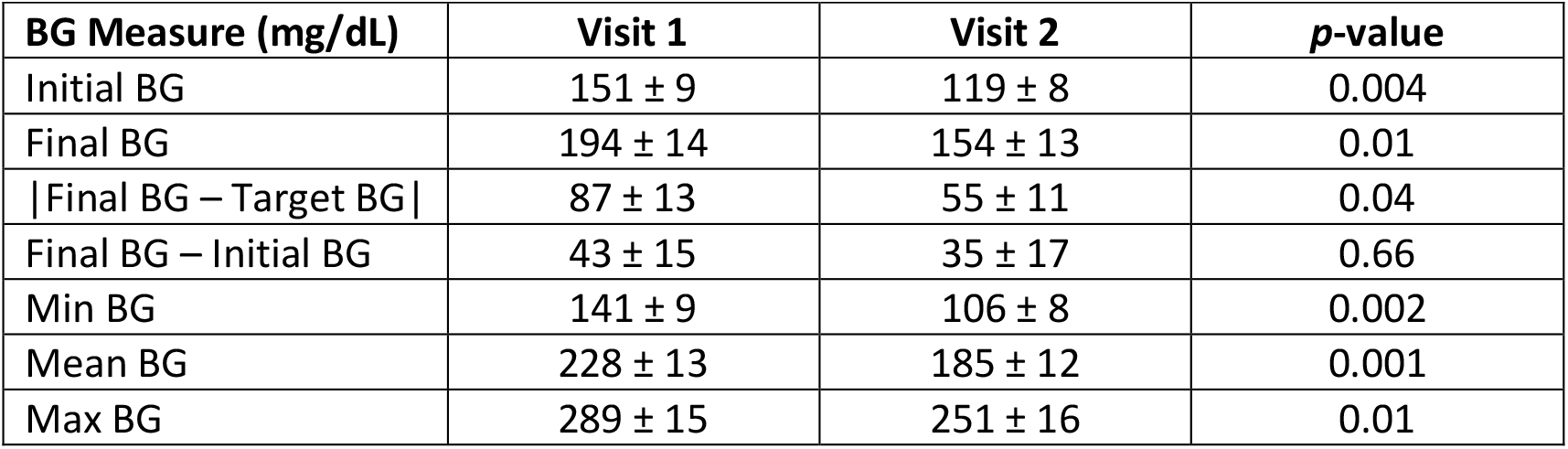
Comparison of various BG measures (mean ± SEM across subjects) between patients’ visit 1 (with the original pump schedule) and visit 2 (with the schedule calibrated using our method). The last column is the two-tailed paired *t*-test *p*-value.

**Figure 1.**
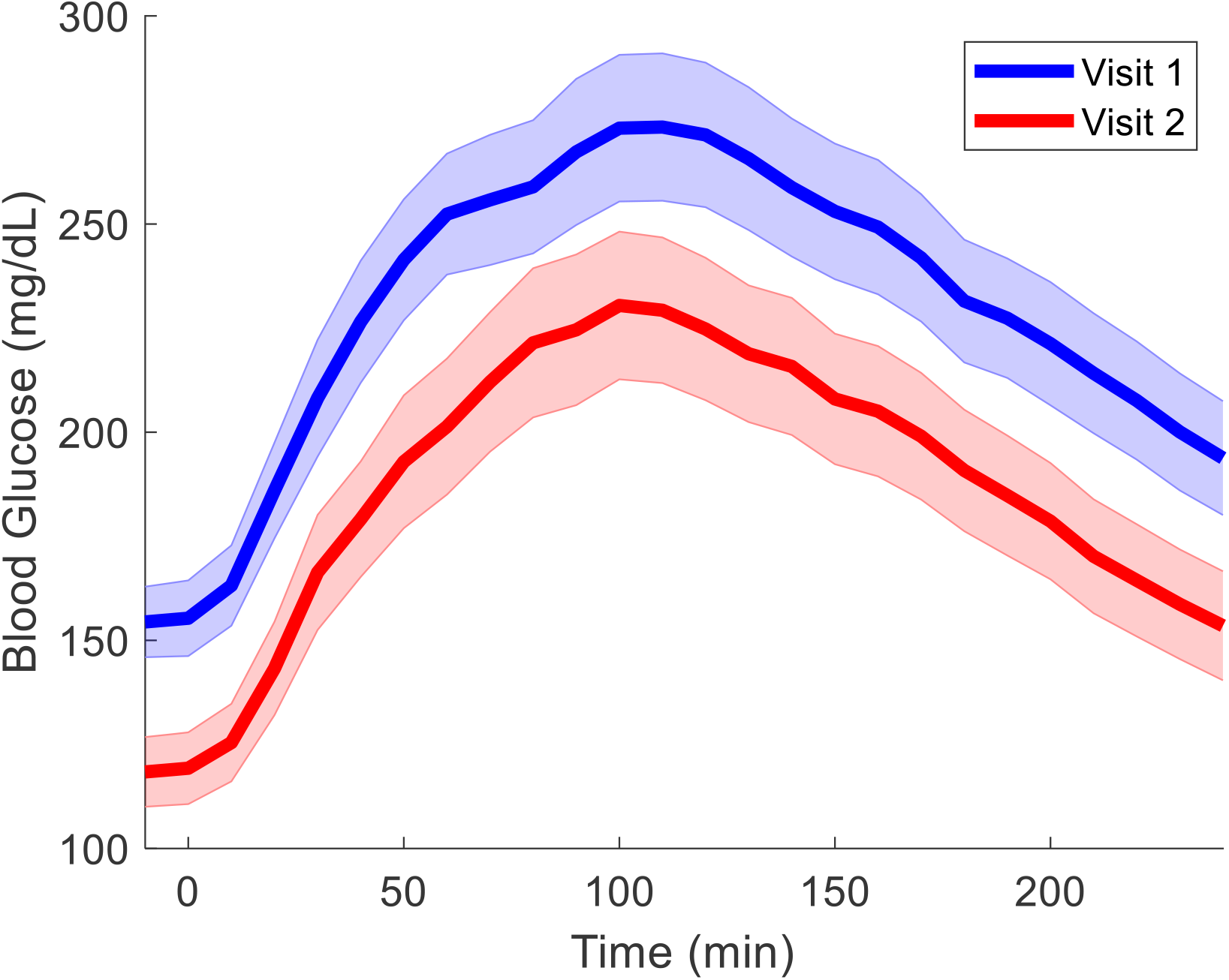
Comparison of the cross-subject average of BG over time between the patients’ visit 1 (blue, with the original pump schedule) and visit 2 (red, with the standard schedule calibrated using our method).

Next, we re-estimated the patients’ parameters simultaneously from visits 1 and 2. The values that we computed for insulin sensitivity (*S*_*I*_) and basal insulin (*k*_2_⁄*S*_*I*_, average of the two values) were significantly correlated with the values originally set on the pump in visit 1 for both insulin sensitivity (r = 0.69, p = 0.0008) and basal insulin (r = 0.54, p = 0.01), across the 20 patients.

We also explored the relationship between the patients’ estimated parameters (7 parameters after averaging the two hepatic glucose rates) and patient data, namely age, sex, weight, and height. We used a two-group two-tailed *t*-test to compare the estimated parameters between sex groups, and used Pearson’s correlation with age, weight, and height. Since 7×4=28 correlations were simultaneously evaluated, we Bonferroni-corrected the *p*-values by multiplying them by 28, which we denote with p_b_. Some of the resulting relationships are plotted in Figure 2. Insulin sensitivity was the estimated parameter most differentiated between sexes; it was higher in females (98 ± 13 mg/dL / u) than in males (41 ± 10 mg/dL / u), which was significant (p = 0.003), albeit not much after Bonferroni correction (p_b_ = 0.08). The next most significant relationship with sex was that of the hepatic glucose rate (*k*_2_), which was higher in females (69 ± 10 mg/dL / h) than in males (33 ± 6 mg/dL / h), with p = 0.005 and p_b_ = 0.15. Among continuous-variable correlations, insulin sensitivity was significantly negatively correlated with body weight (r = -0.67, p = 0.001, p_b_ = 0.037). We then repeated the Pearson tests independently in each sex group, while Bonferroni-correcting with a factor of 7×3×2=42 (for age, weight, and height of the two sex groups). We observed that, in males, the rate of the portion of the plasma insulin that flows to the interstitial fluid (*k*_3_) significantly positively correlated with body weight (r = 0.88, p = 0.0009, p_b_ = 0.036).

**Figure 2.**
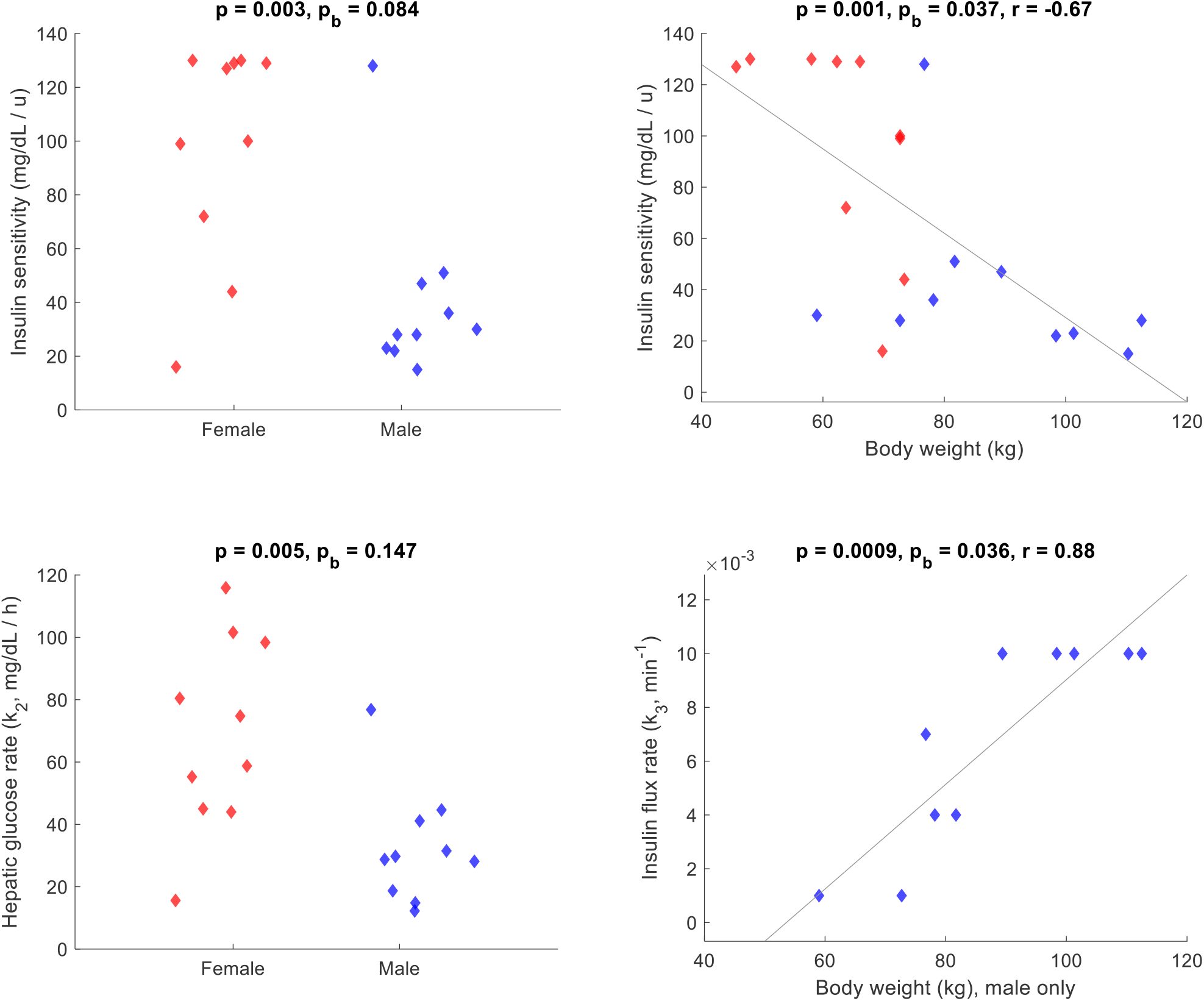
Relationships between some physiological variables estimated from our model (vertical axes) and patient data (horizontal axes). Female and male are represented with red and blue, respectively.

### Optimal insulin delivery

Next, we compared visit 2 (which used a standard schedule with parameters estimated from visit 1) with visit 3 (which used our optimal insulin schedule with parameters estimated simultaneously from visits 1 and 2). We included subjects with equal target BG across the two visits and excluded the first 6 subjects (as we had not yet finalized the protocol for visit 3), resulting in 7 subjects in this comparison.

The results are shown similarly to above in Table 3 and Figure 3. No statistically significant differences were found between the two visits. Most BG values were slightly higher in visit 3 than in visit 2, but this difference might be attributed to the higher initial BG in visit 3.

**Table 3.**
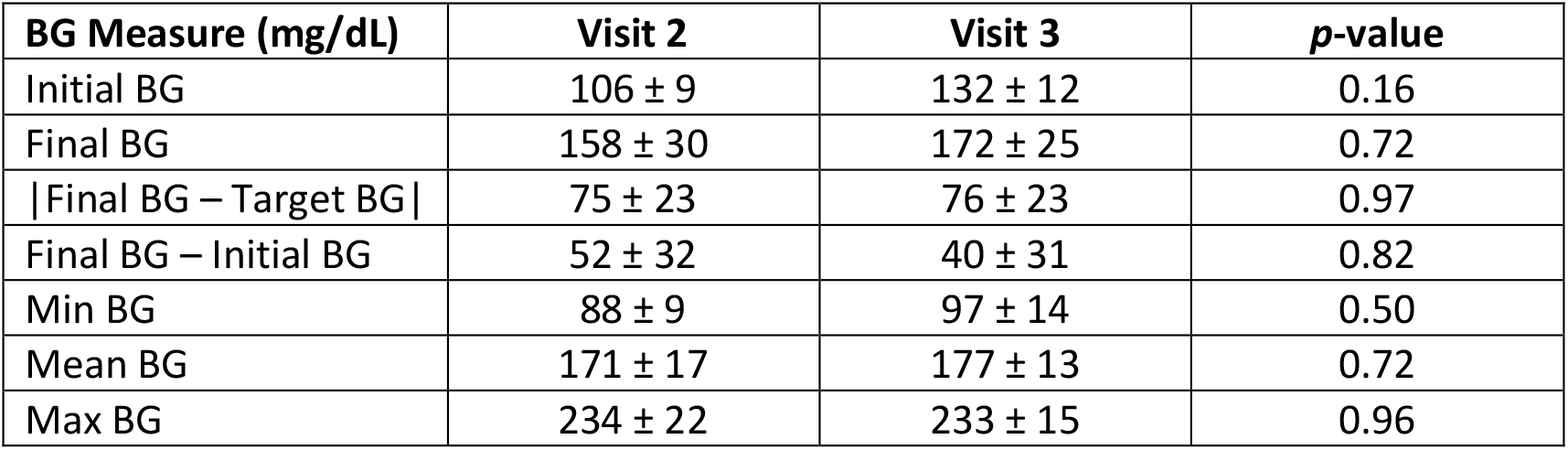
Comparison of various BG measures (mean ± SEM across subjects) between patients’ visit 2 (with the standard schedule) and visit 3 (with our optimal schedule), in both of which physiological parameters had been estimated using our calibration method. The last column is the two-tailed paired *t*-test *p*-value.

**Figure 3.**
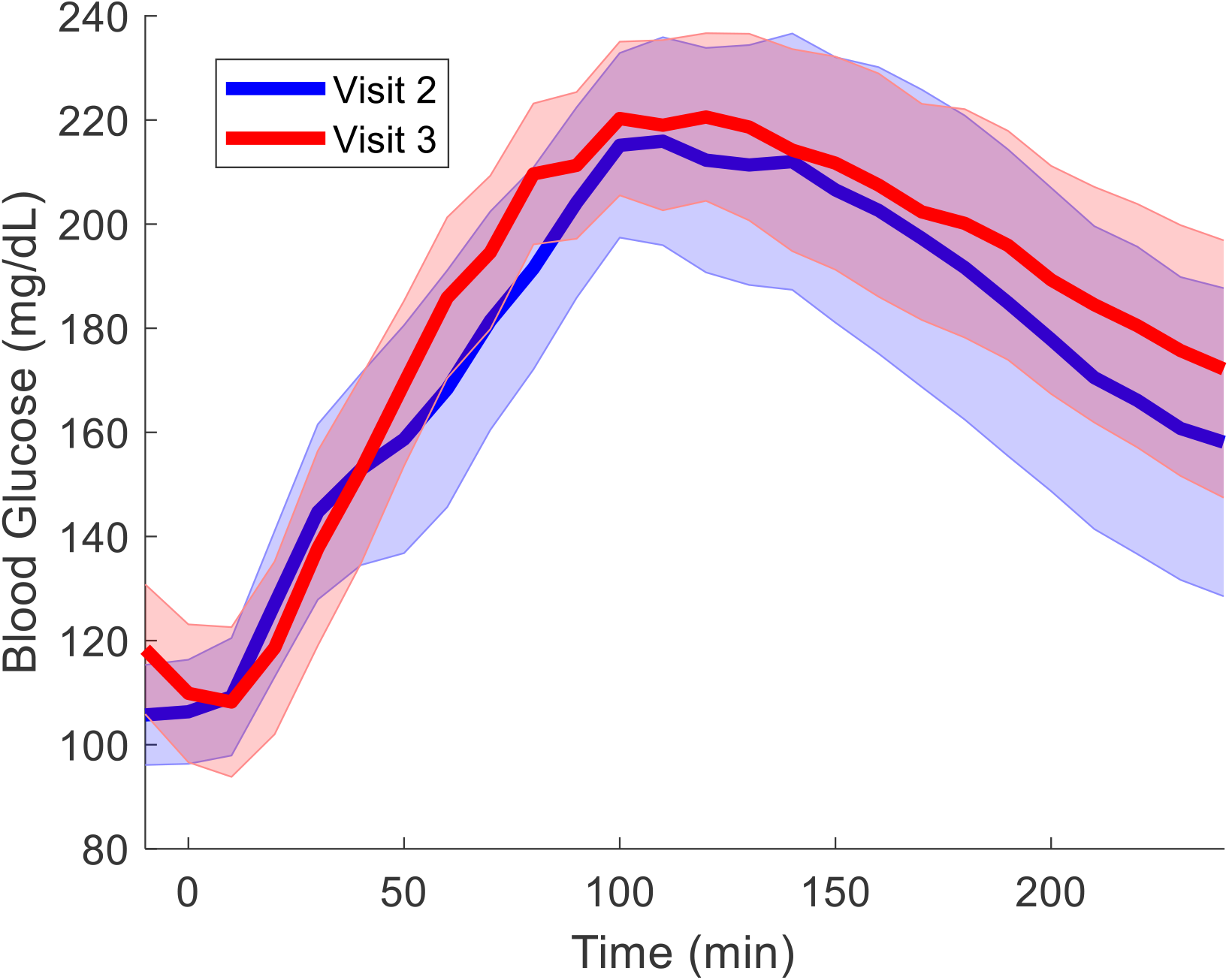
Comparison of the cross-subject average of BG over time between the patients’ visit 2 (blue, with the standard schedule) and visit 3 (red, with our optimal schedule).

Finally, we compared visits 3 and 4 for our last 6 subjects, where the difference was allowing for an earlier pre-bolus in visit 4 (1h) than in visit 3 (35 min). No significant difference in the results of the two visits was found.

Our optimal-scheduling algorithm always recommended turning the basal insulin off (during the *T* = *2h* period where it could optimize the insulin wave) and delivering all that insulin as a single or double pre-bolus. In most cases, it would recommend the pre-bolus to be as early as it was allowed to (35 min and 1h in visits 3 and 4, respectively), except when the initial BG was too close to *G*_*min*_.

## Discussion

We have introduced two computational algorithms, one for insulin pump calibration from a series of BG measurements, and another for insulin delivery optimization. We assessed the performance of these algorithms across 79 BG-measurement sessions, each lasting 4 hours.

A sanity check for the first algorithm was provided through the significant correlation between the parameter values that we estimated and those originally set on the patients’ pumps. A standard insulin schedule based on our estimated values significantly reduced BG from visit 1 to visit 2, highlighting the importance of accurate parameter estimation.

We also observed that the estimated insulin sensitivity was significantly negatively correlated with male sex and body weight. Such relationships have been reported in the literature^38, 39^ and attributed to estrogen^40-42^ and body fat distribution^43-46^, respectively.

Optimal insulin dosing, computed using our model, consistently recommended pre-bolusing all the insulin, likely because subcutaneously administered insulin takes a long time to reach the bloodstream. Essentially, it gathered the basal insulin that would have been given over subsequent hours and delivered it as a few large boluses, primarily to counteract the constraints imposed by subcutaneous insulin kinetics, thereby affecting BG levels more rapidly than is currently possible with the typical basal-bolus model. This is, in fact, a computational derivation of super-bolusing, which has already been suggested in the literature.^33-35^ We did not, however, observe a statistically significant difference between the standard and optimal schedules (visits 2 and 3, in both of which cases, calibration was done with our first method).

Our pilot study has several limitations. First, as we continued learning and recruited more subjects, we occasionally fine-tuned our protocol in the first half of recruitment, especially our optimal scheduling approach (visits 3 and 4). This, along with the fact that the target BG was not consistent across all visits, prevented us from using all our collected data in our comparative analyses, thereby reducing the statistical power. Second, the fact that the initial BG inevitably varied from visit to visit made it less straightforward to interpret our comparative results. The calibration/scheduling strategies juxtaposed here should thus be compared by accounting for the variability in initial BG.

The techniques we have proposed, while used for open-loop control here, will also provide important capabilities for closed-loop systems as CGM technology improves and closed-loop control becomes widely available. Other future work includes incorporating the effects of exercise into our models, with the goal of predicting imminent low BG levels sufficiently far in advance to allow for the ingestion of preventive carbohydrates. We also look forward to conducting a larger study to transition the algorithms into use on an actual pump platform.

## Conclusion

We proposed using models of insulin-glucose kinetics and optimization theory, with the long-term goal of improving the lives of people living with type-1 and insulin-dependent type-2 diabetes. The primary novelty of this project is the use of mathematical modeling and numerical optimization to calibrate insulin pumps and derive insulin delivery schedules that are optimal for maintaining postprandial blood glucose levels within the desired range. We validated our approach on 20 subjects, observing reduced glucose levels following our proposed calibration.

## Data Availability

All Matlab codes in the present study are available upon reasonable request to the authors for research purposes.

## Authorship Confirmation Statement

BF, JLG, IA, and PC contributed to the conceptual design of the study and the writing of the manuscript. NB and JLG recruited the patients and collected the data. IA and BF developed the algorithms and analyzed the data. All authors contributed to the presentation and interpretation of the data and provided substantial feedback on the manuscript.

## Authors’ Disclosure Statement

B. Fischl is a medical advisor to DeepHealth, a company whose medical pursuits focus on brain imaging and measurement technologies. His interests were reviewed and are managed by Massachusetts General Hospital and Mass General Brigham in accordance with their conflict-of-interest policies.

## Funding statement

Support for this research was provided by the National Institute of Diabetes and Digestive and Kidney Diseases (R21DK108277, K01DK101631) of the National Institutes of Health (NIH).

Additional support was provided by the National Institute on Aging (R56AG068261, RF1AG068261, R01AG022381, R01AG016495), the National Institute for Biomedical Imaging and Bioengineering (P41EB015896, R01EB019956, R01EB023281), the National Institute for Neurological Disorders and Stroke (R01NS105820, R01NS083534, U01NS086625), and the BrightFocus Foundation (A2016172S).

Computational resources were provided by NIH Shared Instrumentation Grants (S10RR023401, S10RR019307, S10RR023043, S10RR028832), the O2 High Performance Compute Cluster at Harvard Medical School, and the Enterprise Research Infrastructure & Services at Mass General Brigham.

## Notes

### Author Declarations

This Committee on Human Studies of Joslin Diabetes Center gave ethical approval for this work.

